# Spatial non-equilibrium and distribution dynamic evolution of the development level of national health in China’s provinces

**DOI:** 10.1101/2023.06.15.23291474

**Authors:** Qin Xiao, Haiting Xiao

## Abstract

**Background:** Physical health is the material basis of a country’s socio-economic development, and an in-depth study of the spatial pattern and dynamic evolution of the distribution of national physical fitness in China is of great practical significance for improving national physical fitness.

**Methods:** Based on the national health physique monitoring data of 31 provinces (cities, districts) in China in 2005, 2010 and 2015, the Gini coefficient and its decomposition method and nonparametric density estimation method were used to study the population, males, females, urban and rural nationals in China. Spatial non-equilibrium characteristics and dynamic evolution trend of healthy physique development level.

**Results:** The regional disparities in the development level of health and physique among the general, female, urban and rural nationals in China all show a trend of narrowing at first and then expanding.There is obvious heterogeneity in the regional disparities in the development level of national health and physique within the three regions of East, Middle and West. The distribution of the nuclear density curve of the development level of the Chinese national health constitution showed a dynamic characteristic of evolution from a “spiky, multi-peak” shape to a “broad-peak, single-peak” shape.

**Conclusions:** The low-level provinces have the characteristics of “club convergence”. However, there is a “catch-up effect” between provinces with low levels of rural national health and physique development.

## Introduction

In 2016, the “Healthy China “2030” Planning Outline issued by the Central Committee of the Communist Party of China and the State Council clearly proposed to promote the construction of a healthy China, comprehensively improve the health quality of the Chinese nation, and achieve the coordinated development of people’s health and economic and social development [1]. As an important part of national health, national physique is an important manifestation of the country’s comprehensive national strength, competitiveness, and social civilization and progress [2]. In the important strategic opportunity period of comprehensively promoting the construction of a healthy China, systematically study the spatial characteristics of the development level of Chinese national health and its dynamic evolution trend, which will help improve the relevant policies and measures for promoting the construction of a healthy China and the fairness and accessibility of health, and promote regional national health. Coordinated development and comprehensive improvement of national health and quality of life have important theoretical and practical significance.

In recent years, the research on Chinese national health constitution has become a hot spot of scholars in the field of sports health. Existing research is mainly divided into two categories: one is based on micro-investigation to explore the development level of different groups of national health and its influencing factors. This type of research is mainly based on questionnaires in local areas or some specific groups. Du Faqiang and Fan Jingjing (2014) conducted an ordered multi-class logistic regression analysis on the relationship between their physical fitness levels and influencing factors with 1800 young students as the survey objects [3]. Pan Di et al. (2016) constructed a model of fitness awareness and fitness behavior affecting college students’ physical fitness, and discussed the factors affecting college students’ physical fitness awareness and fitness behavior [4]. Li Kejiang et al. (2019) analyzed the influence of factors such as health literacy, physical exercise, lifestyle, and academic pressure on college students’ physique through health examination and questionnaire survey of students in Chongqing University of Science and Technology [5]. Wang Dan (2018) studied the path dependence of the fit between physical health and survival education of college students [6]. The second is to study the regional distribution of the development level of the national health constitution from the macro level. Such studies are relatively weak. (2017) comprehensively used spatial autocorrelation, Gini coefficient decomposition, stepwise regression and other methods to study the regional differences and social determinants of changes in Chinese national health constitution [7]. Wang Li and Hu Jingchao (2017) used Ordinary Least Squares (OLS) and Geographically Weighted Regression (GWR) models to analyze the spatial distribution characteristics of Chinese nationals’ health and fitness levels [8]. Wei Dexian and Lei Wen (2018) used Exploratory Spatial Data Analysis (ESDA) to study the spatial characteristics and evolutionary trends of the development level of national health in Chinese provinces [9]. Ren Pengbo (2018) conducted a comparative study on the physical health of Hui, Tibetan and Uyghur college students in Northwest China [10].

The above research provides a good reference for this paper, but there are still some shortcomings. On the one hand, based on the availability of data, most of the research at the micro level is aimed at adolescents or college students, ignoring the research on the physical development level of the middle-aged and elderly groups, and it is difficult to provide a full picture of the development level of Chinese national health. At the same time, there is also a lack of comparative research on the development level and differences of urban and rural, male and female national health and physical fitness. On the other hand, based on the limitations of the method, although some scholars have discussed the spatial differences in the development level of Chinese national health at the macro level, they have not been able to further analyze the composition and sources of the differences. At the same time, there is also a lack of analysis from a dynamic perspective The formation and evolution of differences. Based on this, this paper uses the Gini coefficient and its subsample decomposition method to study the spatial disequilibrium degree, composition and source of the development of national health and physique in China’s provinces based on the comprehensive index of national health and physique in 31 provinces in China in 2005, 2010 and 2015., and use the non-parametric density estimation system to analyze the dynamic evolution trend of the spatial disequilibrium of the development level of national health and physique, in order to more comprehensively explore the spatial distribution and dynamic evolution law of the development level of Chinese national health and physique, so as to comprehensively grasp the development of Chinese national health physique. The horizontal spatial distribution and its dynamic evolution trend provide a reference.

## Methods

### Gini coefficient and its decomposition method

Based on the national health physique monitoring data of 31 provinces (cities, districts) in China in 2005, 2010 and 2015, the Gini coefficient and its decomposition method and nonparametric density estimation method were used to study the population, males, females, urban and rural nationals in China.The data underlying this article are available from https://www.sport.gov.cn/. These data were tested by specialized testers organized by the General Administration of Sports of China, all with the consent of the students and their guardians, and the data are publicly available and free of charge.

As one of the classic methods for the study of differences in various fields, the Gini coefficient and its decomposition method can overcome the problems that the Theil index and the traditional Gini coefficient cannot effectively solve the problems of overlapping and overlapping sample data and cannot further analyze the composition of regional differences and their sources (Dagum, 2008). 1997). In this paper, the Gini coefficient and its decomposition method are used to analyze the spatial disequilibrium degree, composition and origin of the development level of Chinese national health. Its specific definition is shown in formula (1):

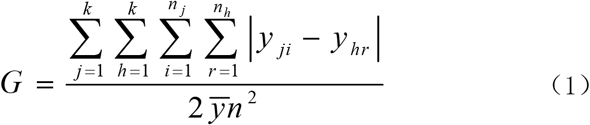

Among them, 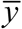 represents the overall level of the development of national health and physique in China, and the value is expressed by the average value of the development level of national health and physique in all provinces (cities and autonomous regions) in the country; *y*_*ji*_ (*yhr*)represents the development level of national health and physique in any province in the *j*(*h*)region, *n*represents the total number of provinces, *k* represents the number of divided regions, and *n*_*j*_(*n*_*h*_)represents the number of provinces in the *j*(*h*) region. In order to further analyze the composition and source of the spatial disequilibrium in the development level of national health and physique in the province, it is first necessary to rank the average development level of national health and physique in the region, as shown in formula (2):

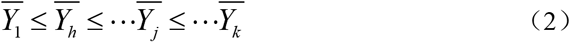

According to the decomposition method of Dagum (1997), G can be further decomposed into the sum of the contribution of intra-regional gaps (*G*_*w*_), the contribution of inter-regional gaps (*G*_*nb*_) and the contribution of hypervariable density (*G*_*t*_), namely:

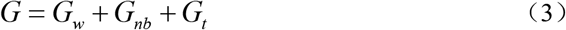

The definitions of 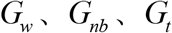 are shown in equations (4)-(6) respectively:

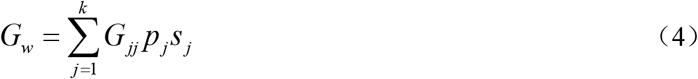

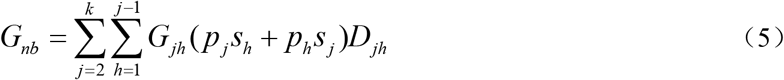

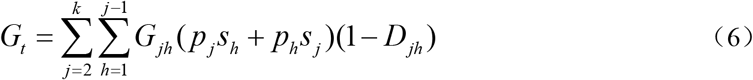

Among them 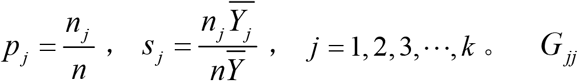 represents the Gini coefficient of the development level of national health in the *j* region, *G*_*jh*_ represents the Gini coefficient of the development level of national health and physique between 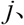 *h* regions, *D*_*jh*_ represents the degree of influence between regions relative to the contribution rate of the national health and fitness development *j*(*h*)level,,the definitions are shown in equations (7)-(9)ο

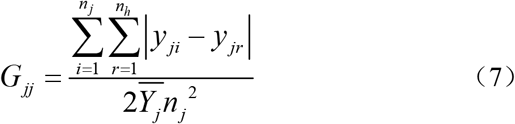

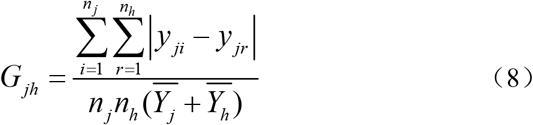

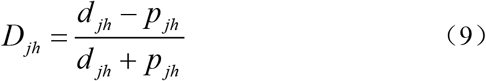

In the formula 9, *d* _*jh*_ is the difference in the contribution rate of the development level of national health and fitness between regions, *p* _*jh*_ is the hypervariable first-order moment, which is defined as formulas (10) and (11) respectively.

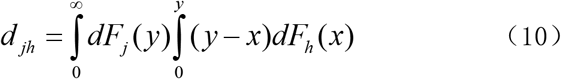

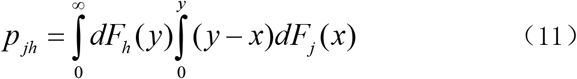

Among them, *F*_*j*_(*F*_*h*_)represents cumulative density distribution function of area *j h*ο

### The Estimation Models of Nonparametric Density *Kernel*

In this paper, kernel density estimation is used to further analyze the dynamic evolution characteristics of provincial national health physique development level. Density estimation is a non-parametric statistical method for fitting the density function of sample distribution in statistical problems feature.

For the dataset{*x*_1_, *x*_2_ … *x*_*n*_},the basic form of the density estimation *Kernal* function is

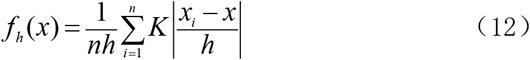

Where is the h bandwidth, N is the number of observations, x is the average value of the observations, *k*(·)representing the kernel density, which is a weighting function or a smooth transition function, usually satisfying:

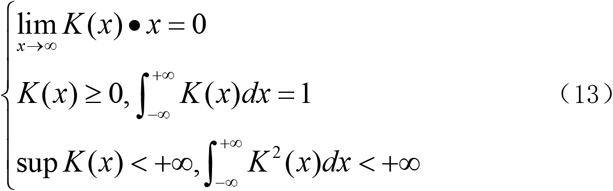

The *Kernel* function mainly includes four types: Gaussian kernel, exponential kernel, triangular kernel and quartic kernel. In this paper, Gaussian kernel is selected to estimate the dynamic evolution law of the development level of national health in China’s provinces. Its function form is:

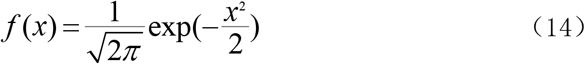

After the function is estimated, the dynamic evolution trend of the variable under investigation can be judged by examining the distribution position, shape and ductility of the variable kernel density estimation curve.

### Data sources and regional definition

This paper uses the comprehensive index of national health and physique in China’s provinces (cities and districts) to measure the development level of national health and physique in each province. The data mainly includes three aspects of body shape, physical function and physical quality, and involves five categories of national health and physical fitness index of 31 provinces (cities, districts), towns, villages, males, and females, which are very representative. Since the survey did not cover regions such as Hong Kong, Macau, and Taiwan, these three regions were not included in the analysis. In order to ensure the comparability of the data, all the data are processed uniformly with the first national health physique monitoring data in 2000 as the base.

Due to the large differences in economy, culture, infrastructure, etc. between regions, this paper further divides the 31 provinces into eastern (including 11 provinces) and central (including 8 provinces) according to the regional division standards of the National Bureau of Statistics), the west (including 12 provinces) three major regions.

## Results

### Visual description of spatial disequilibrium in the development level of national health and fitness in China’s provinces

With the help of ArcGIS software, the spatial distribution map of the overall development level of Chinese national health in 2005 and 2015 was drawn, as shown in Figure 1 and Figure 2. From this, it can be seen that there is a significant spatial disequilibrium in the development level of national health and fitness in China’s provinces. Compared with 2005, in 2015, the number of provinces with a comprehensive index of national health and physique development level above 100 decreased significantly, and the number of provinces between 95 and 100 increased significantly. Although Shanghai was the province with the highest level of national health and physical development in 2005 and 2015, its comprehensive index of national health and physical fitness dropped by 1.67 compared with 2005. Jiangsu ranked second in national health and physical fitness level in 2005 (the comprehensive index was 105.7), in 2015, the national health and physique level dropped significantly (the comprehensive index was 102.92); and Zhejiang ranked second in the country with a comprehensive index of 104.99. In 2005 and 2015, the three provinces of Tibet, Qinghai and Guizhou ranked the bottom three in the country in terms of national health and physical fitness.

**Figure 1.**
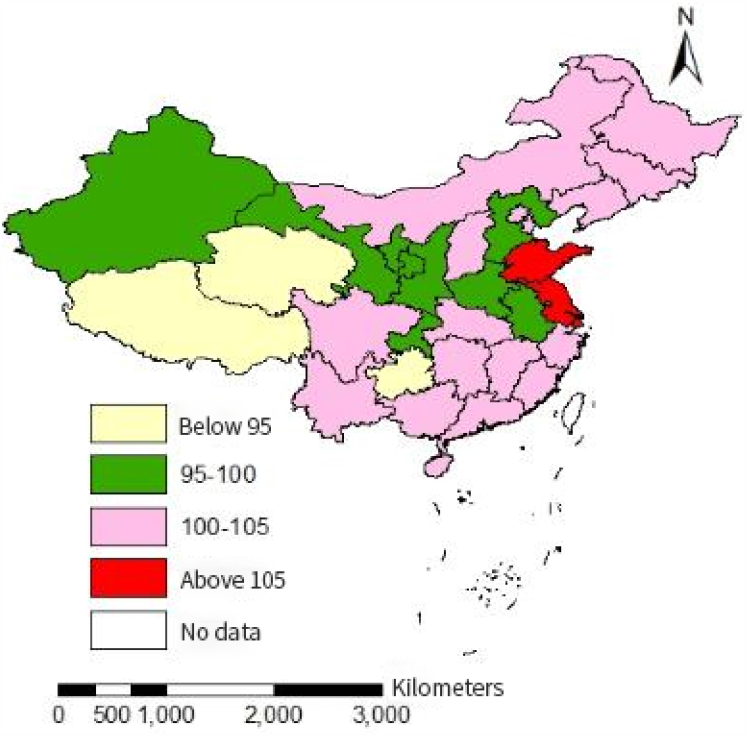
Heat map of regional distribution of the overall level of Chinese national health in 2005

**Figure 2.**
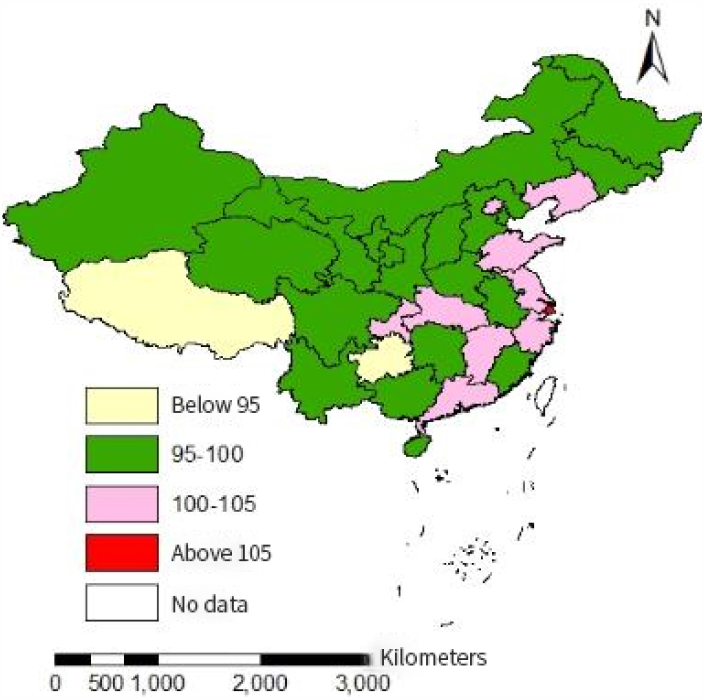
Heat map of regional distribution of the overall level of Chinese national health in 2015

### The composition and source analysis of the spatial disequilibrium in the development level of Chinese national health

In order to further analyze the regional disparities in the spatial distribution of the development level of China’s national health and fitness, its composition, and sources, we used the Gini coefficient and its decomposition method to calculate the overall, male, female, urban, and rural national health in 2005, 2010 and 2015, respectively. The Gini coefficient of the physical development level is further decomposed according to the three major regions of the east, the middle, and the west. The results are shown in Table 1-3.

**Table 1.** Regional Gini coefficients and their decomposition results of the development level of China’s overall national health

| years | overall<br>Coeffi-<br>cient | intra-regional<br>Gini coefficient |  |  | Interregional<br>Gini Coefficient |  |  | Contribution<br>rate (%) |  |  |
| --- | --- | --- | --- | --- | --- | --- | --- | --- | --- | --- |
|  |  | east | Central | west | East-Central | East-West | Central-West | within<br>the<br>area | Interre-<br>gional | Hypervari-<br>able<br>density |
| 2005 | 0.0170 | 0.0127 | 0.0057 | 0.0205 | 0.0115 | 0.0249 | 0.0184 | 0.2715 | 0.6130 | 0.1155 |
| 2010 | 0.0167 | 0.0121 | 0.0094 | 0.0158 | 0.0176 | 0.0230 | 0.0143 | 0.2580 | 0.5444 | 0.1975 |
| 2015 | 0.0177 | 0.0162 | 0.0100 | 0.0118 | 0.0176 | 0.0263 | 0.0147 | 0.2478 | 0.6469 | 0.1053 |

### Regional disparities in the spatial distribution of the overall level of national health and fitness and their sources

Table 1 shows the Gini coefficient and its decomposition results of regional disparities in the overall level of Chinese national health. In 2005, 2010, and 2015, the overall Gini coefficient of the development level of China’s national health and physique showed a trend of first decline and then rising, the lowest in 2010 (0.0167), and 0.07 and 0.1 percentage points higher than that in 2005 and 2010 respectively. It means that during the inspection period, the overall regional gap in the development level of Chinese nationals’ health and physique narrowed first and then expanded. In terms of sub-regions, the results of the Gini coefficient within the region show that the Gini coefficient of the overall level of national health and physique development in the eastern region shows a trend of first decreasing and then increasing, which is consistent with the changing trend of the overall national level; the Gini coefficient of the central region has increased from 2005 The Gini coefficient in the western region gradually decreased from 0.0205 in 2005 to 0.0118 in 2015, showing a downward trend year by year. It shows that there is obvious heterogeneity in the development trend of regional disparities in the level of national health and physique in the three major regions of China during the investigation period. The regional disparity in the development level of national health and physique in the eastern region first narrowed and then expanded, the regional gap in the development of national health and physique in the central region steadily expanded, and the regional gap in the development of national health and physique in the western region decreased year by year. From the perspective of regional disparities, the gap in the development level of national health and physique between the east and the west shows an evolution trend of first narrowing and then expanding, and its Gini coefficient is significantly higher than that of the east-central and central-west, which means that the overall level of national health and physique The east of the regional gap is the largest in the west [12]. From the comparison of East-Central and Central-Western, the regional gap between East-Central in 2005 (0.0115) was significantly lower than that of Central-Western (0.0184), while the regional gap between East-Central in 2010 and 2015 was higher than the Midwest. It shows that the regional disparity growth rate of the development level of national health and physique between the eastern and central regions is higher than that of the central and western regions. From the perspective of the source of the overall regional disparity and its contribution rate, the contribution rate of the inter-regional disparity is stable at more than 54%, and the contribution rate in 2005 and 2015 both exceeded 60%, indicating that the overall regional disparity in the development level of Chinese national health and fitness Mainly due to the gap between regions. The second is the intra-regional gap, whose contribution rate is stable between 24.78% and 27.15%, and generally shows a downward trend over time. The contribution rate of hypervariable density was the lowest, reaching the highest in 2010, at 19.75%, and in 2015, at the lowest, at 10.53%, showing a change characteristic of first increase and then decrease.

### Regional disparities in the spatial distribution of male and female national health and fitness development levels and their sources

Table 2 shows the Gini coefficient and its decomposition results of the regional disparity in the development level of Chinese male and female national health. From the perspective of males, the regional Gini coefficient of the development level of Chinese male national health has increased from 0.0153 in 2005 to 0.0159 in 2015, showing an increasing trend year by year. From the perspective of the intra-regional Gini coefficient, the Gini coefficient of the development level of national health and physique in the eastern and central regions shows an increasing trend year by year, which is consistent with the changing trend of the overall level of men in the country; 0.0182 in 2005 gradually decreased to 0.0105 in 2015, showing a downward trend year by year.

**Table 2.** Regional Gini coefficients of male and female national health and physical development levels and their decomposition

| Category | years | overall<br>coefficient | intra-regional<br>Gini coefficient |  |  | Interregional<br>Gini Coefficient |  |  | Contribution<br>rate (%) |  |  |
| --- | --- | --- | --- | --- | --- | --- | --- | --- | --- | --- | --- |
|  |  |  | east | Central | west | East-Central | East-West | Central-West | within<br>the<br>area | Interre-<br>gional | Hype-<br>rvari-<br>able<br>density |
| male | 2005 | 0.0153 | 0.0104 | 0.0069 | 0.0182 | 0.0104 | 0.0221 | 0.0169 | 0.2717 | 0.6022 | 0.1261 |
|  | 2010 | 0.0154 | 0.0125 | 0.0100 | 0.0136 | 0.0170 | 0.0203 | 0.0127 | 0.2681 | 0.5153 | 0.2167 |
|  | 2015 | 0.0159 | 0.0157 | 0.0103 | 0.0105 | 0.0162 | 0.0225 | 0.0133 | 0.2628 | 0.5985 | 0.1387 |
| female | 2005 | 0.0195 | 0.0154 | 0.0069 | 0.0232 | 0.0138 | 0.0279 | 0.0207 | 0.2774 | 0.5816 | 0.1410 |
|  | 2010 | 0.0185 | 0.0143 | 0.0102 | 0.0177 | 0.0186 | 0.0258 | 0.0160 | 0.2633 | 0.5508 | 0.1860 |
|  | 2015 | 0.0199 | 0.0177 | 0.0105 | 0.0145 | 0.0196 | 0.0298 | 0.0167 | 0.2481 | 0.6516 | 0.1004 |

From the comparison of the Gini coefficients of male national health and physical development levels in the three regions, in 2005 and 2010, the Gini coefficient in the western region was the largest, followed by the eastern region and the smallest in the central region; in 2015, the Gini coefficient in the eastern region was the largest, followed by the western region and the smallest in the central region. Judging from the Gini coefficient between regions, the gap in the development level of male national health and physique between the eastern and western regions showed an evolution trend of first decreasing and then increasing, and the gap was always higher than that of the eastern-central and central-western regions during the investigation period [13]. The Gini coefficient between the eastern and central regions showed a trend of first increasing and then decreasing, reaching a maximum value in 2010; the Gini coefficient between the central and western regions generally showed a downward trend. From the perspective of the source of the overall regional disparity and its contribution rate, the contribution rate of the regional disparity has been stable at more than 51%, indicating that the overall regional disparity in the development level of Chinese male national health is still dominated by the regional disparity. The second is the intra-regional gap, whose contribution rate is stable between 24.81% and 27.74%, and generally shows a downward trend over time. The contribution rate of hypervariable density is the lowest. During the investigation period, the highest value appeared in 2010, which was 21.67%, and the lowest value appeared in 2015, which was 13.87%.

From the perspective of women, the overall Gini coefficient of the regional gap and the Gini coefficient of the eastern region generally showed a trend of first decreasing and then increasing. From the perspective of the inter-regional Gini coefficient, the Gini coefficient between the eastern and western regions is still the largest. During the investigation period, its variation trend is similar to that of the central and western regions, showing a trend of first decreasing and then increasing. The Gini coefficient shows an increasing trend year by year. From the perspective of the source of the overall regional disparity and its contribution rate, the contribution rate of the inter-regional disparity is still the largest, accounting for more than 55% over the years, followed by the intra-regional disparity, but the contribution rate of the intra-regional disparity shows a downward trend year by year. The contribution rate of the over-editing density is the lowest, and the overall evolution trend during the investigation period showed an evolution trend of first increasing and then decreasing [14]. Compared with men, the overall regional disparity in the development level of female national health and physical fitness is significantly larger than that of men.

### Regional disparities in the spatial distribution of urban and rural national health and fitness development levels and their sources

Table 3 shows the Gini coefficient and its decomposition results of the regional disparity in the development level of national health in urban and rural areas in China. From the perspective of cities and towns, the overall Gini coefficient of the regions with the development level of urban national health was basically stable at around 0.0185 in 2005 and 2010 and rose sharply to 0.0238 in 2015. The Gini coefficients within the eastern and central regions increased steadily year by year, from 0.0152 and 0.0075 in 2005 to 0.0374 and 0.0109 in 2015, respectively. The western region showed a gradual decline, from 0.0210 in 2005 to 0.0109 in 2015. From the perspective of the Gini coefficient between regions, the Gini coefficient between the eastern and western regions is still the largest, and generally shows a trend of first decreasing and then increasing. The Gini coefficient between the peaks reached 0.0193 in 2005, and was basically stable at around 0.0145 in 2005 and 2010. From the perspective of the source of the overall urban regional gap and its contribution rate, the contribution of the regional gap in 2005 and 2010 dominated, accounting for 58.26% and 44.86%, respectively. In 2015, the contribution of hypervariable density dominated, reaching 46.32%. On the whole, the contribution of the inter-regional gap showed a significant downward trend, while the contribution of the intra-regional and hypervariable density showed a steady increase, and the growth rate of the contribution of the super-density density was significantly greater than that of the intra-regional gap. From the perspective of the development level of rural national health and physique, the overall Gini coefficient of regional disparity and the internal Gini coefficient of the eastern region showed a trend of first decline and then increase, the central region showed a slight upward trend, and the western region showed a significant decline year by year. And by 2015, the internal disparity in the eastern region surpassed that in the western region [15]. It means that the evolution trend of the regional disparity in the development level of national health and physique in the eastern, central and western regions is first narrowing and then widening, slightly expanding year by year, and greatly expanding year by year. From the perspective of regional gaps, the regional gap between the east-west region is still the largest, and with the passage of time, the regional gaps between the east-west and east-central regions first increased and then narrowed, and the middle-Regional differences between the western regions are just the opposite. By 2015, the regional gap between the eastern and central regions surpassed that of the central and western regions, ranking second.

**Table 3.**
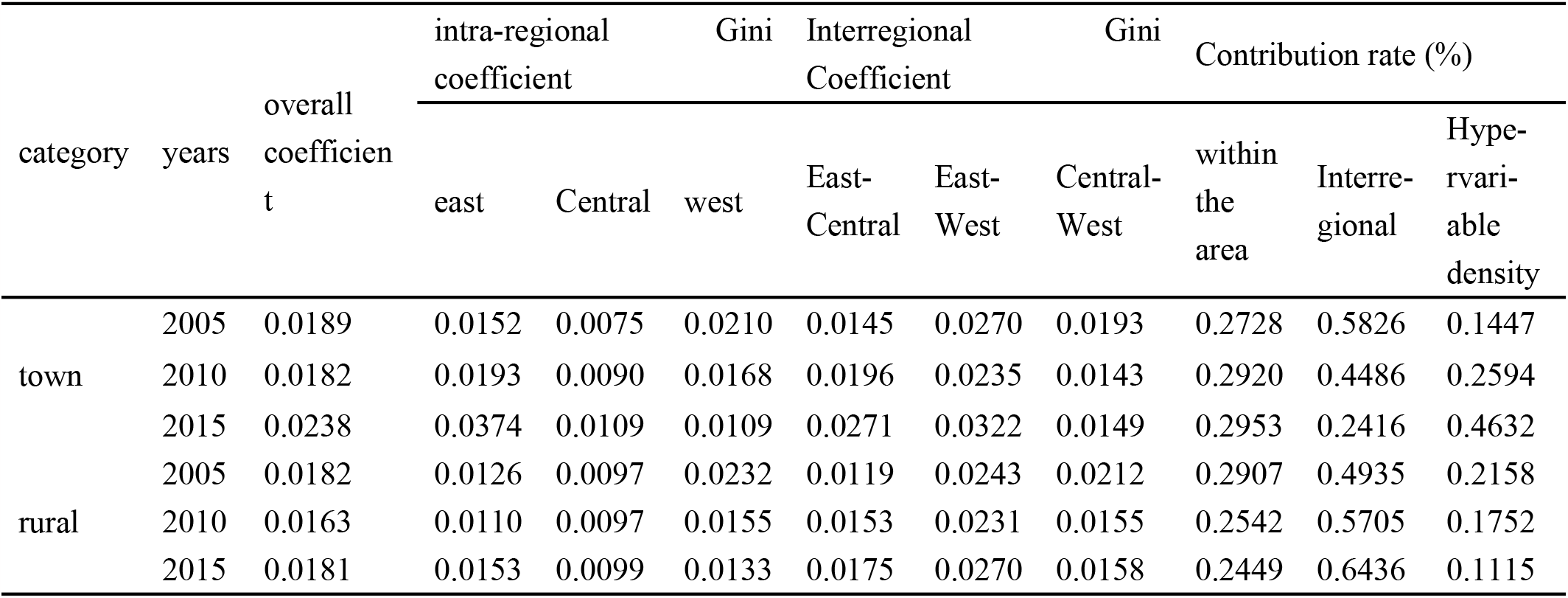
Regional Gini coefficient and its decomposition of the development level of urban and rural national health

## Discussion

### The dynamic evolution of the spatial distribution of the development level of Chinese national health

From the perspective of relative disparity, the above analysis analyzes the degree of spatial disequilibrium in the development level of Chinese national health and fitness, its composition, and sources. In the following, we further analyze the dynamic evolution trend of the spatial distribution of Chinese national health physique development level from the perspective of absolute disparity and spatial distribution form using a *Kernel*-density estimation method.

### *Kernel* estimation of the dynamic evolution of the overall national health and fitness level

Figure 3 shows the distribution of the nuclear density curve of the spatial distribution of the overall national health and fitness development level in China from 2005 to 2015. From this, it can be seen that the distribution curve of nuclear density in 2005 presents a two-tailed multi-peak shape, indicating that the development level of national health and physique in some provinces is concentrated to a higher level, and the development level of national health and physique in some provinces is concentrated to a lower level, showing a “high”. The higher the higher, the lower the lower”. By 2010 and 2015, the distribution curve changed significantly: the height of the peak decreased significantly, the width of the peak increased, and the right-wing further extended slightly, showing the characteristics of evolution from a “spiky, multi-peak” shape to a “broad peak, single-peak” shape. It shows that while the overall development level of Chinese nationals’ health and physique has improved slightly during the inspection period, regional disparities have also further widened.

**Figure 3.**
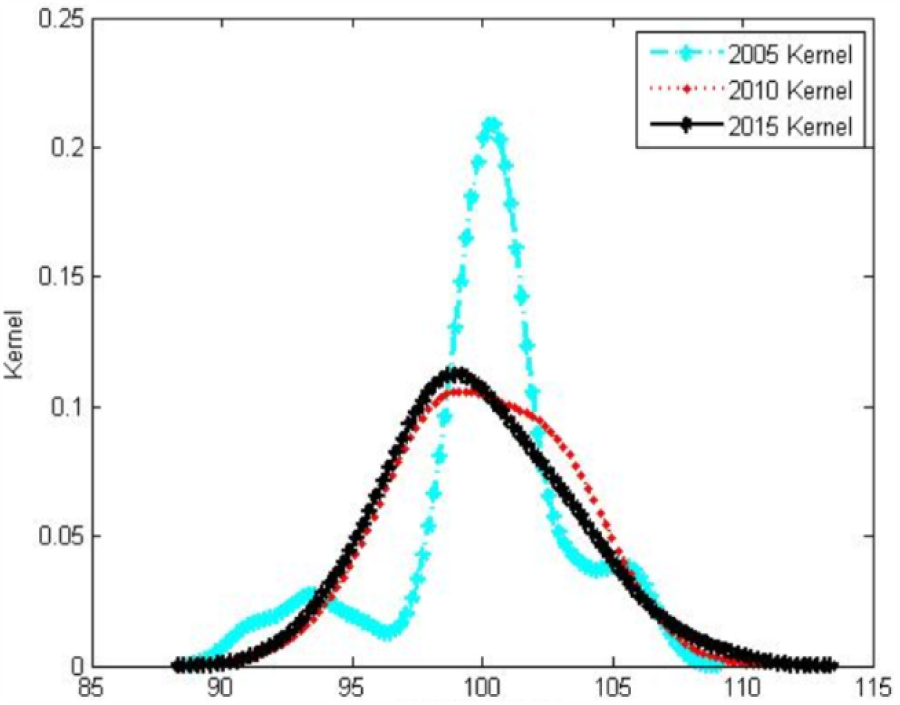
Kernel density estimation diagram for the dynamic evolution of the overall national health physique level

Figures 4 and 5 are the distributions of the kernel density estimation curves of the development level of Chinese men’s and women’s national health and physique from 2005 to 2015, respectively. It can be seen from this that the dynamic evolution trend of the spatial distribution of male and female national health physique development levels has a strong similarity. From 2005 to 2010, the nuclear density estimation curves of the national health physique development level of both males and females showed that the peak value of the peak continued to decrease, the right tail slightly extended, the kurtosis gradually widened, and the evolution from a multi-peak shape to a single-peak shape was characterized. However, the nuclear density curve did not change much from 2010 to 2015 and remained basically stable. It shows that the regional disparity in the development level of male and female national health and physique has further increased on the whole. Among them, because the provinces with low levels of national health and physique of men and women show a certain “catch-up effect”, the polarization of low-level national health physiques of different genders gradually disappears, while the provinces with higher development levels of national health physiques The physical level has also been further improved. From the point of view of the peak position, the distribution curve of males remains basically unchanged, while the distribution curve of females shifts slightly to the left, indicating that the average level of Chinese male national health and physique has remained basically stable as a whole, while the average female national health physique has shown a slight increase. decline. From the comparison of the peaks, the peak value of the nuclear density curve of males far exceeds the level of 0.1, while the peak value of the nuclear density curve of females basically remains at the level of about 0.1, indicating that the absolute regional gap in the development level of male national health is generally smaller than women.

**Figure 4.**
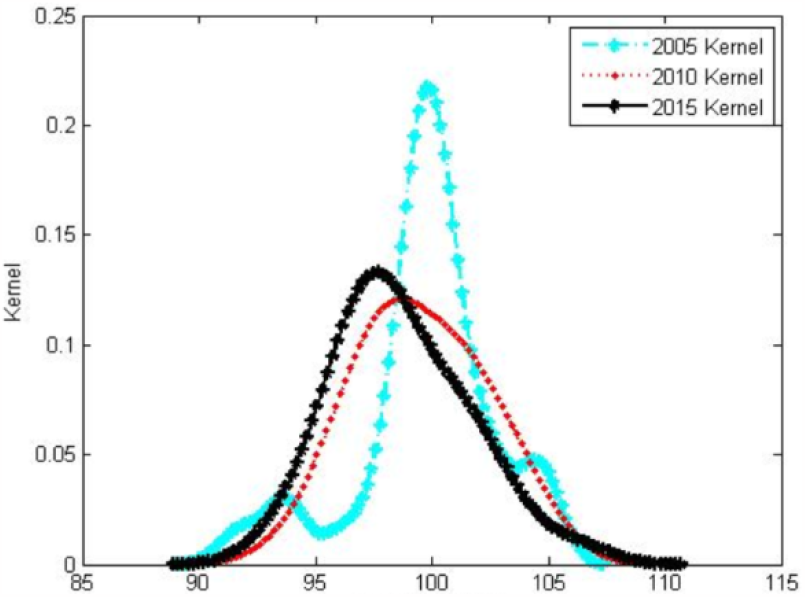
Kernel density estimation diagram for the dynamic evolution of female national health physique level

**Figure 5.**
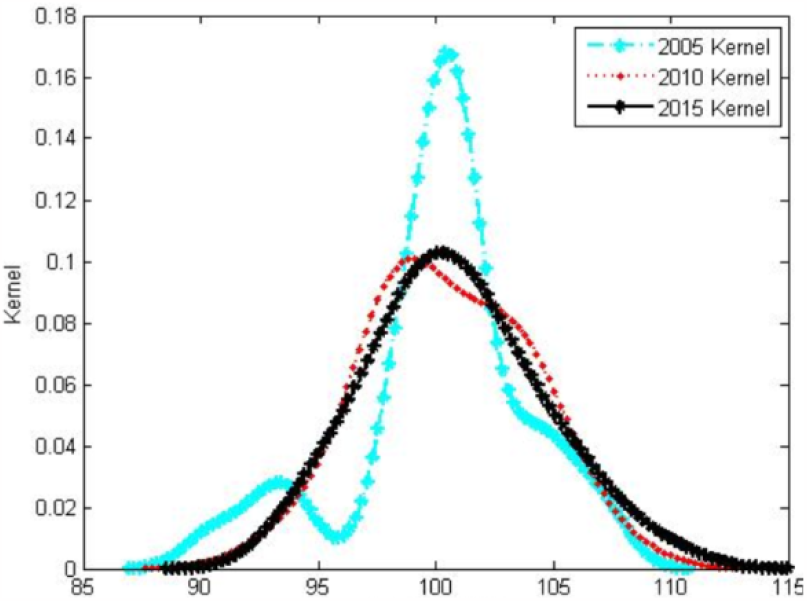
Kernel density estimation diagram for the dynamic evolution of male national health physique level

Figures 6 and 7 are the distributions of the kernel density estimation curves of the development levels in urban and rural areas of China from 2005-2015, respectively. It can be seen from the development level of national health in urban and rural areas that there is obvious heterogeneity in the dynamic evolution trend of the spatial distribution of the development level of national health and fitness in urban and rural areas in China. As far as towns are concerned, the distribution of nuclear density curves in 2010 and 2015 did not change much, but there was a big difference compared with 2005. With the passage of time, the kurtosis of the curve decreases and shifts slightly to the left, the right tail changes from an obvious single peak to a less obvious double peak, the left tail wave peak is further obvious and shifted to the left, and the left tail tip as a whole is greatly shifted to the left. Extend left. It shows that the regional disparity in the development level of national health and physique in China’s cities and towns has generally further increased, and some provinces with relatively low national health and physique levels are concentrated at lower levels, showing the characteristics of low-level “club convergence”; while high-level physique The trend of provincial concentration to a higher level has gradually eased. From the rural point of view, compared with 2005, the peaks of the curves in 2010 and 2015 decreased, the width of the peaks increased, and the tip of the right tail slightly extended outward, and the curves changed from multi-peak to single-peak. It shows that with the passage of time, the polarization of the development level of rural national health in China gradually disappears, and there is a “catch-up effect” in the physical development level of low-level provinces. The development trend of rural national health in different provinces tends to be Consistent.

**Figure 6.**
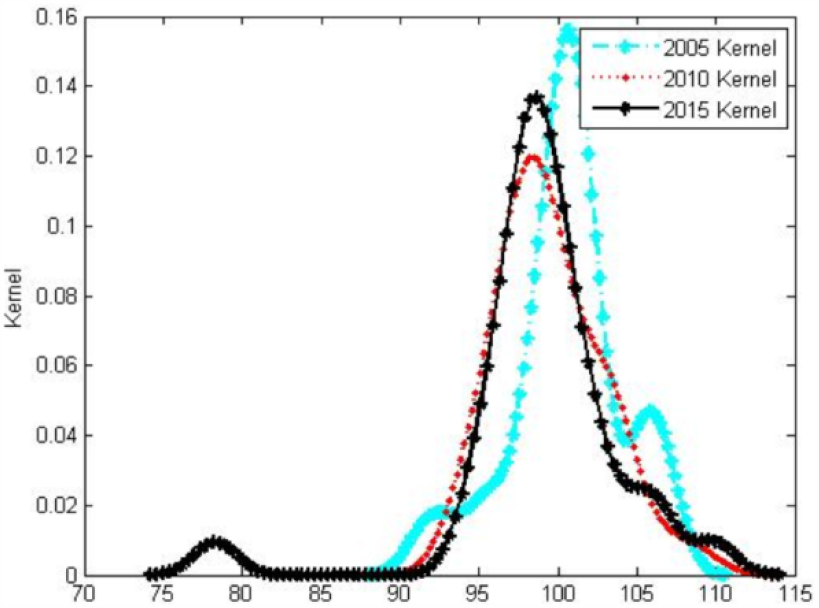
Kernel density estimation for the dynamic evolution of urban national health physique level

**Figure 7.**
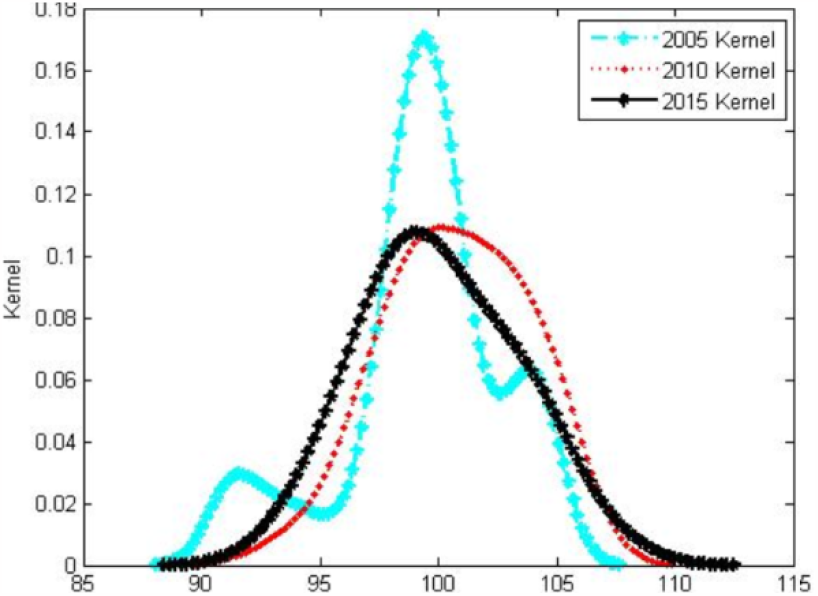
Kernel density estimation diagram diagram for the dynamic evolution of rural national health physique level

## Conclusions

Based on the national health physique monitoring data of 31 provinces (cities, districts) in China in 2005, 2010, and 2015, this paper uses the Gini coefficient and its decomposition method and nonparametric density estimation method to study the overall, male, and female, urban and rural areas in China. Spatial non-equilibrium and dynamic evolution characteristics of the development level of national health constitution. The research results show that: (1) The regional disparities in the development level of health and physique among the general population, women, and urban and rural nationals in China all show a trend of first narrowing and then expanding, and the regional disparity in the development level of male national health and physique is increasing year by year. (2) From the perspective of intra-regional disparities, there is obvious heterogeneity in the regional disparities in the development level of national health and physique within the three regions. The regional disparity in the overall development level of national health and physique in the eastern, central, and western regions showed a trend of first decline and then rising, steadily rising, and declining year by year. In terms of gender, the regional gap in the development level of male national health and physique in the eastern region shows an increase year by year, while the female shows an evolution trend of first decline and then rise; The West is just the opposite. In terms of urban and rural areas, the regional gap in the development level of urban citizens’ health and physique in the eastern and central regions has expanded year by year, while the western region has shown a decline year by year. The regional disparity in the overall development level of the rural national health constitution and the regional disparity in the eastern region showed a trend of first decline and then increase, the central region showed a slight upward trend, and the western region showed a sharp decline year by year. (3) Inter-regional disparity is the main source of spatial imbalance in the development of Chinese national health and fitness, followed by intra-regional disparity, with the smallest cause of imbalance caused by hyper-variable density. The regional disparity in the development level of national health and physique among regions is most obvious between the east and west regions. (4) There is significant heterogeneity in the dynamic evolution trend of the spatial distribution of the development level of the Chinese national health constitution. With the passage of time, the nuclear density curve of the spatial distribution of the development level of Chinese national health and physique showed a dynamic characteristic of evolution from a “spike-peak, multi-peak” shape to a “broad-peak, single-peak” shape. Specifically, while the overall development level of national health and physique has improved slightly, the absolute gap between regions has also further expanded. There are strong similarities in the dynamic evolution trends of the spatial distribution of male and female national health physique development levels. The development level of male national health and physical fitness is generally lower than that of females. Provinces with low levels of national health and physique development of both males and females showed a certain “catch-up effect”, which made the low-level polarization of national health physiques of different genders gradually disappear, while provinces with higher national health physique development levels showed a certain “catch-up effect”. The level has also been further improved, and the final manifestation is that the regional gap in the development of male and female national health and physique has further expanded. There is obvious heterogeneity in the dynamic evolution trend of the spatial distribution of the development level of national health in urban and rural areas in China. With the passage of time, the absolute regional disparity in the development level of urban national health and physique has further increased. The trend of concentrating on a higher level gradually eased. With the passage of time, the polarization of the development level of rural people’s health and physique has gradually disappeared, and there is a “catch-up effect” in the level of physique development between provinces with low levels of the physique. Consistent.

Based on the above conclusions, this paper puts forward the following policy recommendations: First, based on the large regional disparity in the development level of national health and physique, it is necessary to formulate a coordinated development policy of regional national health and physique under the overall framework to narrow the development level of national health and physique between regions. the gap, and comprehensively improve the level of national health and physique. Second, based on the fact that the current level of Chinese national health and fitness has not increased significantly and regional disparities are widening, it is necessary to vigorously improve the public service system for national fitness and effectively improve and enhance the health and fitness of the people. At the same time, differentiated design and reasonable Allocate health resources, steadily promote the construction of a unified medical security system covering the whole population and gradually realize that each citizen will not be affected by differences in urban and rural household registration, gender, and provincial economic and social development levels. Access to medical and health resources and security levels, and effectively promote national health equity.

## Data Availability

ll relevant data are within the manuscript and its Supporting Information files.

## Author contributions

The manuscript was conceived and written by QX, HTX is respon sible for the organization and statistics of the data, QX and HTX revised it. All authors h ave contributed to its writing and approved its publication.

## Acknowledgments

We are grateful to the General Administration of Sports of China for providing us with the dataset. The dataset was made publicly available to us.

## Funding

Project supported by the Ministry of Education of Humanities and Social Science project,China(Grant No. 22YJC890038), the Social Sciences Foundation of Hunan Province, China (Grant No.17YBX008), Philosophy and social science planning project of Guang dong Province(Grant No. GD20CTY10), The key research and development Plan Project in Hunan Province(2020SK2104).

## Competing interests

None declared.

## Ethical approval

Approval for the use of 2005, 2010, and 2015 data was granted by the Hunan Normal University Biomedical Research Ethics Review Committee (2022/388).

## Data availability

The data underlying this article are available from https://www.sport.gov.cn/.

## Notes

### Competing Interest Statement

The authors have declared no competing interest.

### Funding Statement

yes

